# The global prevalence and genetic spectrum of primary carnitine deficiency

**DOI:** 10.1101/2024.05.29.24308100

**Authors:** Liu Sun, Hang-Jing Wu

## Abstract

**Background:** Primary carnitine deficiency (PCD) is an autosomal recessive rare disorder of carnitine cycle and carnitine transport caused by mutations in the SLC22A5 gene. The prevalence of PCD is unclear. This study aimed to estimate the carrier frequency and genetic prevalence of PCD using Genome Aggregation Database (gnomAD) data.

**Methods:** The pathogenicity of SLC22A5 variants was interpreted according to the American College of Medical Genetics and Genomics (ACMG) standards and guidelines. The minor allele frequency (MAF) of SLC22A5 gene disease-causing variants in 807,162 unique individuals was examined to estimate the global prevalence of PCD in five major ethnicities: African (afr), Admixed American (amr), East Asian (eas), Non-Finnish European (nfe) and South Asian (sas). The global and population-specific carrier frequencies and genetic prevalence of PCD were calculated using the Hardy–Weinberg equation.

**Results:** In total, 195 pathogenic/likely pathogenic variants (PV/LPV) were identified according to ACMG standards and guidelines. The global carrier frequency and genetic prevalence of PCD were 1/88 and 1/31,260, respectively.

**Conclusions:** The prevalence of PCD is estimated to be 1/30,000 globally, with a range of between 1/20,000 and 1/70,000 depending on ethnicity.

## Introduction

Systemic primary carnitine deficiency (OMIM: 212140, ORPHA: 158, GARD: 5104) is a rare inborn error of fatty acid metabolism [1]. It presents with a broad spectrum of clinical signs and symptoms, with cardiac symptoms (predominantly cardiomyopathy) being the most prevalent. Neurological, hepatic, and metabolic symptoms occurred. symptoms in PCD predominantly develop in early childhood, however, adult onset of symptoms can occur, patients suffered a severe event without any preceding symptom. Newborn screening (NBS) can detect most cases of PCD, with both newborns and mothers of newborns remaining asymptomatic [2]. The diagnosis can be suspected on newborn screening, it is established through genetic and/or functional (carnitine transporter activity) testing [3]. Primary carnitine deficiency can be treated with L-carnitine supplementation [4].

PCD is an autosomal recessive disorder which caused by homozygous or compound heterozygous pathogenic mutations in the SLC22A5 gene [5–7] that encodes the plasma membrane sodium-dependent high affinity carnitine transporter OCTN2 [8]. OCTN2 is necessary for L-carnitine transport across the plasma membrane and L-carnitine is necessary for transporting long chain fatty acids into the mitochondria for fatty acid β-oxidation. The SLC22A5 gene, located on chromosome 5q31.1, spans 10 exons and comprises 25,903 base pairs, encoding OCTN2 protein of 557 amino acids. more than 200 *SLC22A5* variants have been identified in patients with PCD since 1999 [7, 9–11].

primary carnitine deficiency (PCD) exact prevalence is unknown and varies depending on ethnicity. The incidence is 1/10,000-1/20,000 in China through newborn screening [12–14], 1/20,000 - 1/70,000 newborns in Europe and the USA [1] while the estimated incidence in Japan is 1/40,000 births [11]. The prevalence of PCD in the Faroe Islands is 1/297, which is the highest reported in the world [15].

As high-throughput sequencing technology has evolved, re-evaluation guidelines for interpreting and classifying the pathogenicity of identified variants have been implemented. In addition, large-scale population databases have become widely available and can be used for assessment of genetic variants in rare diseases. In fact, for several rare diseases, there is evidence that these databases have improved the interpretation and classification of variants in patients with monogenic disease and allowed better prediction of which variants are likely to cause disease. The Genome Aggregation Database (gnomAD) is a resource developed by an international coalition of investigators, with the goal of aggregating and harmonizing both exome and genome sequencing data from a wide variety of large-scale sequencing projects. The v4 data set (GRCh38) spans 730,947 exome sequences and 76,215 whole-genome sequences from unrelated individuals of diverse ancestries.

We attempted to obtain a more reliable estimate of the global prevalence and genetic spectrum of PCD from the Genome Aggregation Database (gnomAD) dataset using a well-established pipeline [16, 17]. Additionally, we aimed to generate a curated machine learning training dataset of *SLC22A5* variants for pathogenicity classification and interpretation.

## Methods

### Simulation single nucleotide variants and missense variants of SLC22A5 gene

To simulate all single nucleotide variants (SNVs) in the SLC22A5 gene, restrict the SNVs to be generated to coding sequence (CDS) from the Matched Annotation from NCBI and EMBL-EBI (MANE) transcript and corresponding protein [18] using the Variation Simulation Python package (https://github.com/liu-sun/VariationSimulation) with Human Genome Variation Society (HGVS) nomenclature [19], and then translate HGVS notation to all possible variant IDs (refSNP, SPDI, VCF, *etc.*) using Python client for Ensembl REST API (https://pypi.org/project/ensemblrestpy/). Simulated missense, nonsense and synonymous variants were included in the subsequent variant annotation and curation.

all missense (SNVs and MNVs) variants in SLC22A5 gene coding sequence (CDS) were simulated using Variation Simulation python package with state-of-the-art of variant prediction score: EVE [20], AlphaMissense [21], ESM1b [22], CADD [23] and PrimateAI-3D [24].

### Identification and annotation of previously reported *SLC22A5* disease-causing variants

To evaluate the genomic spectrum of PCD, a comprehensive literature retrieve was performed to identify all previously reported disease-causing SLC22A5 gene variants. Searches were conducted in the literature database PubMed, Scopus and Web of Science using the following combinations of retrieval terms: primary carnitine deficiency (PCD), carnitine transporter defect (CTD), carnitine uptake deficiency (CUD), SLC22A5, OCTN2, mutations, variants, variations and mutants.

Two authors screened publications according to inclusion and exclusion criteria: original case reports and newborn screening (NBS) reporting disease-causing variants of the SLC22A5 gene were included, and variants in title, abstract, full-text, tables, figures, or supplementary material were extracted. Non-English-language case reports, articles, reviews, comments, editorials, letters, *etc.*, and cell-based assays and animal model studies were excluded. Common SLC22A5 polymorphisms associated with common traits in genome-wide association studies were also excluded.

Reported *SLC22A5* variants were also identified from LitVar [25], Ensembl Variation [26] and ClinVar [27]. Prediction and functional annotation for all *SLC22A5* nonsynonymous single-nucleotide variants were compiled using Ensembl Variant Effect Predictor (VEP).

Predicting splicing consequence for SLC22A5 synonymous single-nucleotide variants using SpliceAI [28] and Pangolin [29].

### Identification and prediction of novel SLC22A5 loss of function variants

The gnomAD database (https://gnomad.broadinstitute.org/) was searched for novel loss of function (LoF) SLC22A5 variants that had not yet been reported, and protein-truncating variants (PTVs) were examined (frameshifts, stop codons, initiator codons, splice donors and splice acceptors).

### Interpretation of the pathogenicity of SLC22A5 variants

The pathogenicity of variants was interpreted following the Clinical Interpretation of Sequence Variants protocol [30]. The pathogenicity of all missense, synonymous, in-frame indel and nonsense, frameshift and splicing SLC22A5 variants was classified according to Standards and Guidelines of the American College of Medical Genetics and Genomics (ACMG) [31] and ClinGen Sequence Variant Interpretation Working Group (SVI) recommendation [32–38] with the ClinGen Variant Curation Interface (VCI) implement [39].

For missense, in-frame indel and synonymous variants, we performed additional literature retrieval to curate *in vitro* or *in vivo* functional studies supportive of functional effect on SLC22A5 missense/synonymous/in-frame variants. Variants reported in a peer-reviewed journal were labeled with PS3 level evidence and classified as likely pathogenic if variants with function less than 20% of WT OCTN2 function with respect to carnitine transport [40, 41].

Variants classified as pathogenic and likely pathogenic were included, and variants classified as benign, likely benign, or uncertain significance were excluded, respectively. Pathogenic/likely pathogenic variants were included in the subsequent carrier frequency and prevalence calculation.

### Annotation of variants with minor allele frequencies (MAFs)

four consequence category *SLC22A5* variants: predicted loss-of-function (pLoF) variants, missense/inframe indel variants, synonymous variants and other variants data were directly downloaded from gnomAD browser. Variants were subsequent manually filtered out, which flagged by the loss-of-function transcript effect estimator (LOFTEE), such as variants near the end of transcripts and in non-canonical splice sites and multi-nucleotide variants (MNVs).

The canonical SLC22A5 transcript (NM_003060.4 / ENST00000245407.8, 3,277 bp) and protein (NP_003051.1 / ENSP00000245407.3, 557 aa) were selected by Matched Annotation from NCBI and EMBL-EBI (MANE) [18].

The minor allele frequency (MAF) of pathogenic/likely pathogenic *SLC22A5* variants in the gnomAD population for the following ethnic groups: African/African American (AFR), American Admixed/Latino (AMR), East Asian (EAS), Non-Finish European (NFE) and South Asian (SAS).

### Carrier frequency and genetic prevalence calculation

The genetic prevalence and carrier frequency of PCD were calculated based on the Hardy–Weinberg equation. For a monogenic autosomal recessive disorder, the genetic prevalence is given by [1−∏_i_(1−q_i_)]^2^. This is based on the theory of probability, where q_i_ stands for each likely pathogenic/pathogenic variant minor allele frequency (MAF). The genetic prevalence was approximately equal to (∑q_i_)^2^, and the carrier frequency was 2(1−∑q_i_) ∑q_i_≈2∑q_i_. The disease prevalence was estimated by utilizing the observed allele frequency of a likely pathogenic/pathogenic variant in the gnomAD v4.1 database as the direct estimator for q_i_. The disease prevalence can be estimated by the following equation [1−∏_i_(1−AC_i_/AN_i_)]^2^≈∑AF_i_^2^. here, the AC is the allele count, the AN is allele number and the AF is allele frequency [16, 17].

The Python statistics package statsmodels and scientific computing package NumPy were employed for calculating the 95% confidence interval (95% CI) for the binomial proportion of carrier frequency and the genetic prevalence with the Clopper–Pearson interval. Graphics were plotted using the python graphic package seaborn and matplotlib [16, 17].

## Results

### Simulation of SLC22A5 single nucleotide variants and missense variants

single nucleotide variants (SNVs) in SLC22A5 gene coding sequence were generated 5013 variants, including 3633 missense variants, 162 nonsense variants, 1218 synonymous variants. Missense variants in SLC22A5 gene coding sequence were generated 10583 variants, including 5013 SNVs and 5570 multi-nucleotide variants (MNVs). All single nucleotide variants and multi-nucleotide variants are listed in the Science Data Bank (ScienceDB) online data repository. three MNVs c.35_36delinsAT (p.Gly12Asp), c.56_57delinsCT (p.Arg19Pro) and c.1324_1325delinsAT (p.Ala442Ile) were previously reported [41–45].

### Identification of *SLC22A5* disease-causing variants

Comprehensive retrieval of PCD disease-causing variants resulted in the identification of 2209 variants in the gnomAD v4.1 database, including 108 pLoF variants, 802 missense/in-frame indel variants, 273 synonymous variants and 1026 other variants. After filter, remaining 94 pLoF variants. For in-frame, synonymous and other variants, we pick reported variants: c.453G>A (p.Val151=) [41, 46] and c.-149G>A (rs57262206) [47, 48], p.Phe23del [49] and p.Leu394del [41], respectively. All disease-causing variant pipelines are shown in Fig. 1.

**Figure 1.**
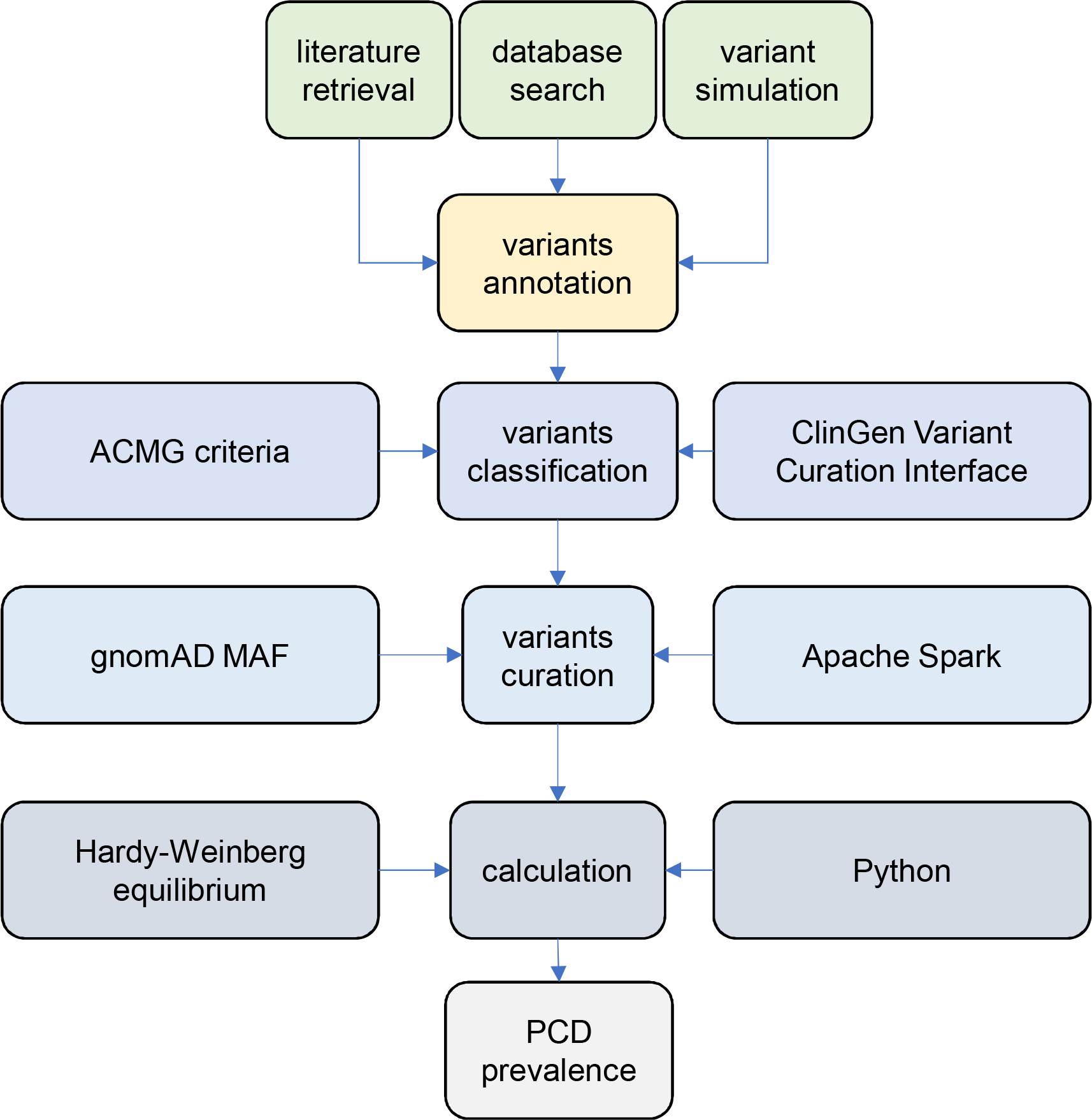
flowchart of curation and classification of SLC22A5 pathogenic/likely pathogenic variants and prevalence calculation.

195 disease-causing variants in the *SLC22A5* gene, including 98 missense variants, 2 in-frame variants, 1 UTR variant and 94 protein-truncating variants (PTVs), are classified as pathogenic/likely pathogenic according to the Standards and guidelines of the American College of Medical Genetics and Genomics and the Association for Molecular Pathology (missense variants are shown in Table 1; all variants are listed in the Science Data Bank (ScienceDB) online data repository).

**Table 1.**
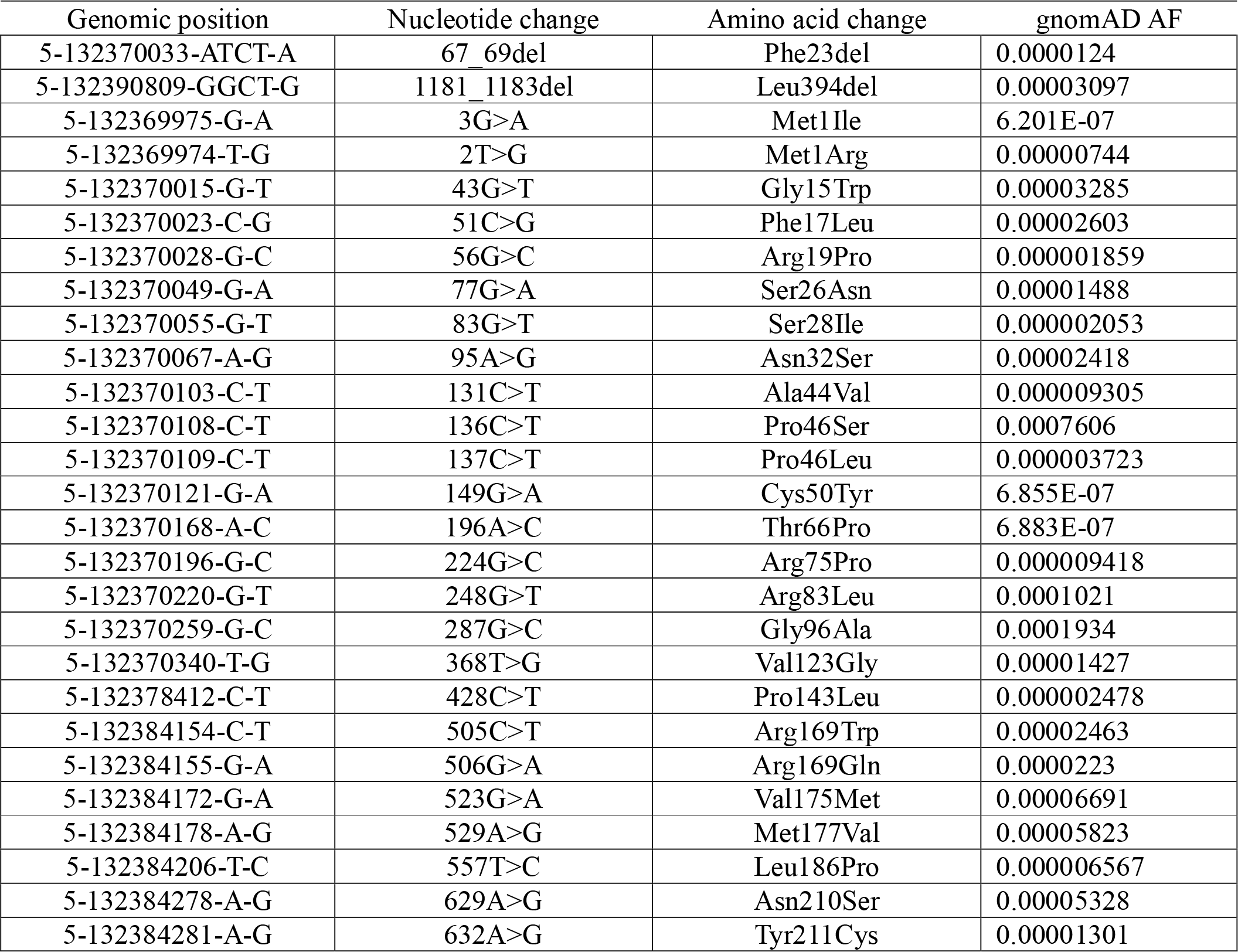
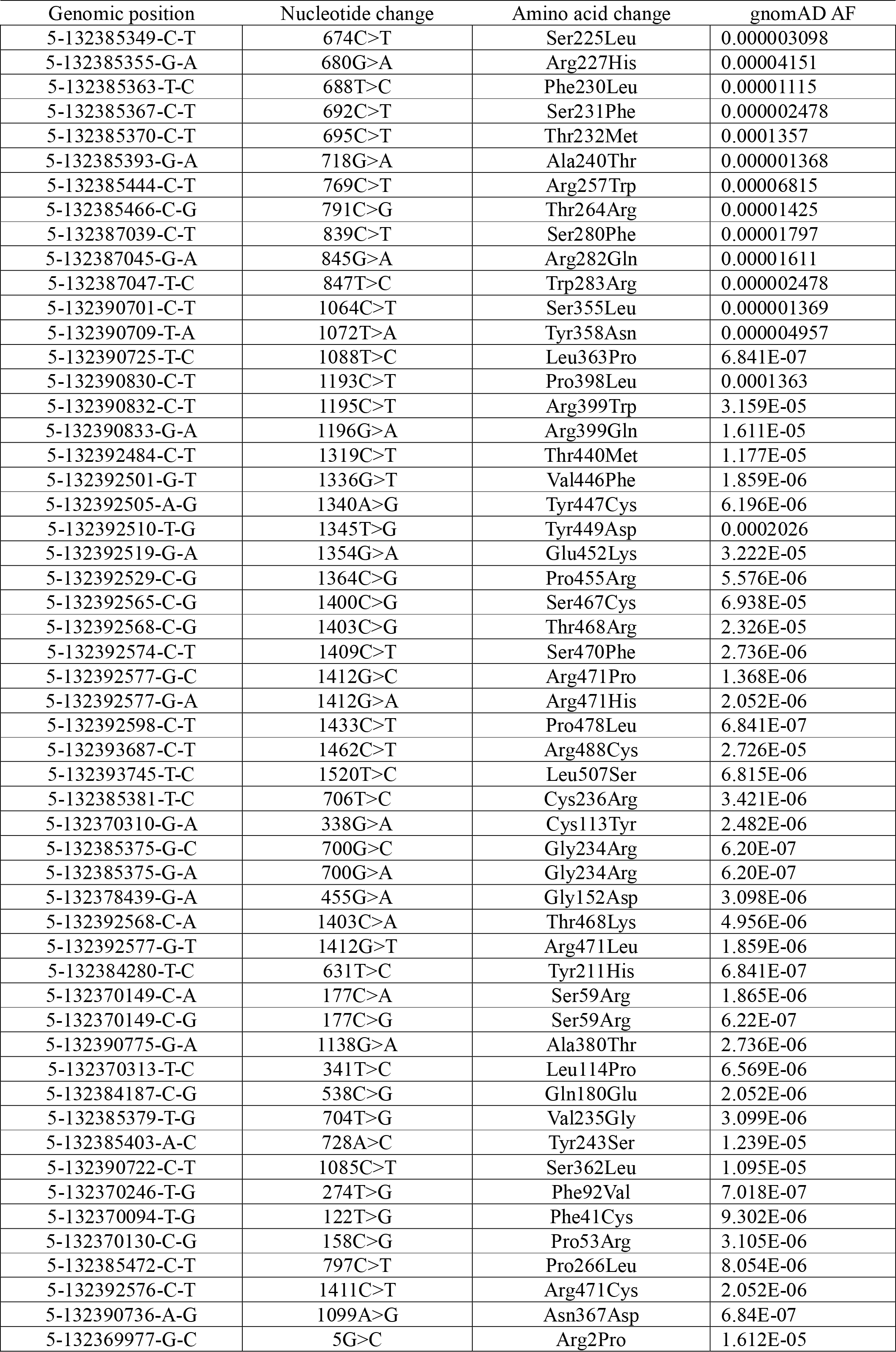
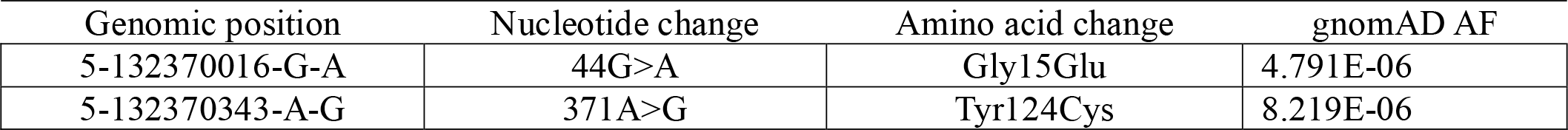
SLC22A5 pathogenic/likely pathogenic missense variants present in gnomAD with global minor allele frequencies.

| Genomic position | Nucleotide change | Amino acid change | gnomAD AF |
| --- | --- | --- | --- |
| 5-132370033-ATCT-A | 67_69del | Phe23del | 0.0000124 |
| 5-132390809-GGCT-G | 1181_1183del | Leu394del | 0.00003097 |
| 5-132369975-G-A | 3G>A | Met1Ile | 6.201E-07 |
| 5-132369974-T-G | 2T>G | Met1Arg | 0.00000744 |
| 5-132370015-G-T | 43G>T | Gly15Trp | 0.00003285 |
| 5-132370023-C-G | 51C>G | Phe17Leu | 0.00002603 |
| 5-132370028-G-C | 56G>C | Arg19Pro | 0.000001859 |
| 5-132370049-G-A | 77G>A | Ser26Asn | 0.00001488 |
| 5-132370055-G-T | 83G>T | Ser28Ile | 0.000002053 |
| 5-132370067-A-G | 95A>G | Asn32Ser | 0.00002418 |
| 5-132370103-C-T | 131C>T | Ala44Val | 0.000009305 |
| 5-132370108-C-T | 136C>T | Pro46Ser | 0.0007606 |
| 5-132370109-C-T | 137C>T | Pro46Leu | 0.000003723 |
| 5-132370121-G-A | 149G>A | Cys50Tyr | 6.855E-07 |
| 5-132370168-A-C | 196A>C | Thr66Pro | 6.883E-07 |
| 5-132370196-G-C | 224G>C | Arg75Pro | 0.000009418 |
| 5-132370220-G-T | 248G>T | Arg83Leu | 0.0001021 |
| 5-132370259-G-C | 287G>C | Gly96Ala | 0.0001934 |
| 5-132370340-T-G | 368T>G | Val123Gly | 0.00001427 |
| 5-132378412-C-T | 428C>T | Pro143Leu | 0.000002478 |
| 5-132384154-C-T | 505C>T | Arg169Trp | 0.00002463 |
| 5-132384155-G-A | 506G>A | Arg169Gln | 0.0000223 |
| 5-132384172-G-A | 523G>A | Val175Met | 0.00006691 |
| 5-132384178-A-G | 529A>G | Met177Val | 0.00005823 |
| 5-132384206-T-C | 557T>C | Leu186Pro | 0.000006567 |
| 5-132384278-A-G | 629A>G | Asn210Ser | 0.00005328 |
| 5-132384281-A-G | 632A>G | Tyr211Cys | 0.00001301 |
| 5-132385349-C-T | 674C>T | Ser225Leu | 0.000003098 |
| 5-132385355-G-A | 680G>A | Arg227His | 0.00004151 |
| 5-132385363-T-C | 688T>C | Phe230Leu | 0.00001115 |
| 5-132385367-C-T | 692C>T | Ser231Phe | 0.000002478 |
| 5-132385370-C-T | 695C>T | Thr232Met | 0.0001357 |
| 5-132385393-G-A | 718G>A | Ala240Thr | 0.000001368 |
| 5-132385444-C-T | 769C>T | Arg257Trp | 0.00006815 |
| 5-132385466-C-G | 791C>G | Thr264Arg | 0.00001425 |
| 5-132387039-C-T | 839C>T | Ser280Phe | 0.00001797 |
| 5-132387045-G-A | 845G>A | Arg282Gln | 0.00001611 |
| 5-132387047-T-C | 847T>C | Trp283Arg | 0.000002478 |
| 5-132390701-C-T | 1064C>T | Ser355Leu | 0.000001369 |
| 5-132390709-T-A | 1072T>A | Tyr358Asn | 0.000004957 |
| 5-132390725-T-C | 1088T>C | Leu363Pro | 6.841E-07 |
| 5-132390830-C-T | 1193C>T | Pro398Leu | 0.0001363 |
| 5-132390832-C-T | 1195C>T | Arg399Trp | 3.159E-05 |
| 5-132390833-G-A | 1196G>A | Arg399Gln | 1.611E-05 |
| 5-132392484-C-T | 1319C>T | Thr440Met | 1.177E-05 |
| 5-132392501-G-T | 1336G>T | Val446Phe | 1.859E-06 |
| 5-132392505-A-G | 1340A>G | Tyr447Cys | 6.196E-06 |
| 5-132392510-T-G | 1345T>G | Tyr449Asp | 0.0002026 |
| 5-132392519-G-A | 1354G>A | Glu452Lys | 3.222E-05 |
| 5-132392529-C-G | 1364C>G | Pro455Arg | 5.576E-06 |
| 5-132392565-C-G | 1400C>G | Ser467Cys | 6.938E-05 |
| 5-132392568-C-G | 1403C>G | Thr468Arg | 2.326E-05 |
| 5-132392574-C-T | 1409C>T | Ser470Phe | 2.736E-06 |
| 5-132392577-G-C | 1412G>C | Arg471Pro | 1.368E-06 |
| 5-132392577-G-A | 1412G>A | Arg471His | 2.052E-06 |
| 5-132392598-C-T | 1433C>T | Pro478Leu | 6.841E-07 |
| 5-132393687-C-T | 1462C>T | Arg488Cys | 2.726E-05 |
| 5-132393745-T-C | 1520T>C | Leu507Ser | 6.815E-06 |
| 5-132385381-T-C | 706T>C | Cys236Arg | 3.421E-06 |
| 5-132370310-G-A | 338G>A | Cys113Tyr | 2.482E-06 |
| 5-132385375-G-C | 700G>C | Gly234Arg | 6.20E-07 |
| 5-132385375-G-A | 700G>A | Gly234Arg | 6.20E-07 |
| 5-132378439-G-A | 455G>A | Gly152Asp | 3.098E-06 |
| 5-132392568-C-A | 1403C>A | Thr468Lys | 4.956E-06 |
| 5-132392577-G-T | 1412G>T | Arg471Leu | 1.859E-06 |
| 5-132384280-T-C | 631T>C | Tyr211His | 6.841E-07 |
| 5-132370149-C-A | 177C>A | Ser59Arg | 1.865E-06 |
| 5-132370149-C-G | 177C>G | Ser59Arg | 6.22E-07 |
| 5-132390775-G-A | 1138G>A | Ala380Thr | 2.736E-06 |
| 5-132370313-T-C | 341T>C | Leu114Pro | 6.569E-06 |
| 5-132384187-C-G | 538C>G | Gln180Glu | 2.052E-06 |
| 5-132385379-T-G | 704T>G | Val235Gly | 3.099E-06 |
| 5-132385403-A-C | 728A>C | Tyr243Ser | 1.239E-05 |
| 5-132390722-C-T | 1085C>T | Ser362Leu | 1.095E-05 |
| 5-132370246-T-G | 274T>G | Phe92Val | 7.018E-07 |
| 5-132370094-T-G | 122T>G | Phe41Cys | 9.302E-06 |
| 5-132370130-C-G | 158C>G | Pro53Arg | 3.105E-06 |
| 5-132385472-C-T | 797C>T | Pro266Leu | 8.054E-06 |
| 5-132392576-C-T | 1411C>T | Arg471Cys | 2.052E-06 |
| 5-132390736-A-G | 1099A>G | Asn367Asp | 6.84E-07 |
| 5-132369977-G-C | 5G>C | Arg2Pro | 1.612E-05 |
| 5-132370016-G-A | 44G>A | Gly15Glu | 4.791E-06 |
| 5-132370343-A-G | 371A>G | Tyr124Cys | 8.219E-06 |

**Table 2.**
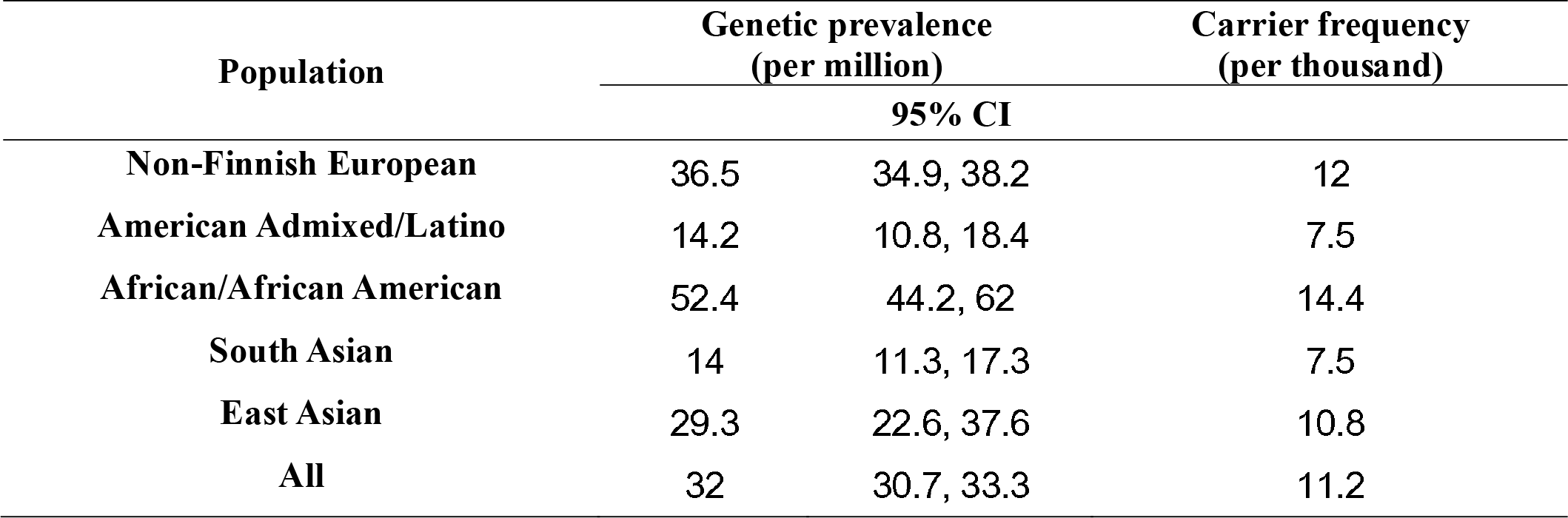
Estimated prevalence and carrier frequency among ethnicities and ancestries.

gnomAD allele frequencies were available for 178/195 disease-causing diseases. Including 81 missense, 2 in-frame, 1 UTR variants and 94 pLoF variants. Pooling of the allele frequencies of these variants resulted in a global allele frequency of 0.005655974, which is equivalent to a prevalence of 3.2 per million population (95% confidence interval (95% CI): [3.07, 3.33]). Five major populations had distinct prevalence (Fig 2 and Table 1) with a range of between 1/20,000 and 1/70,000 depending on ethnicity.

**Fig. 2.**
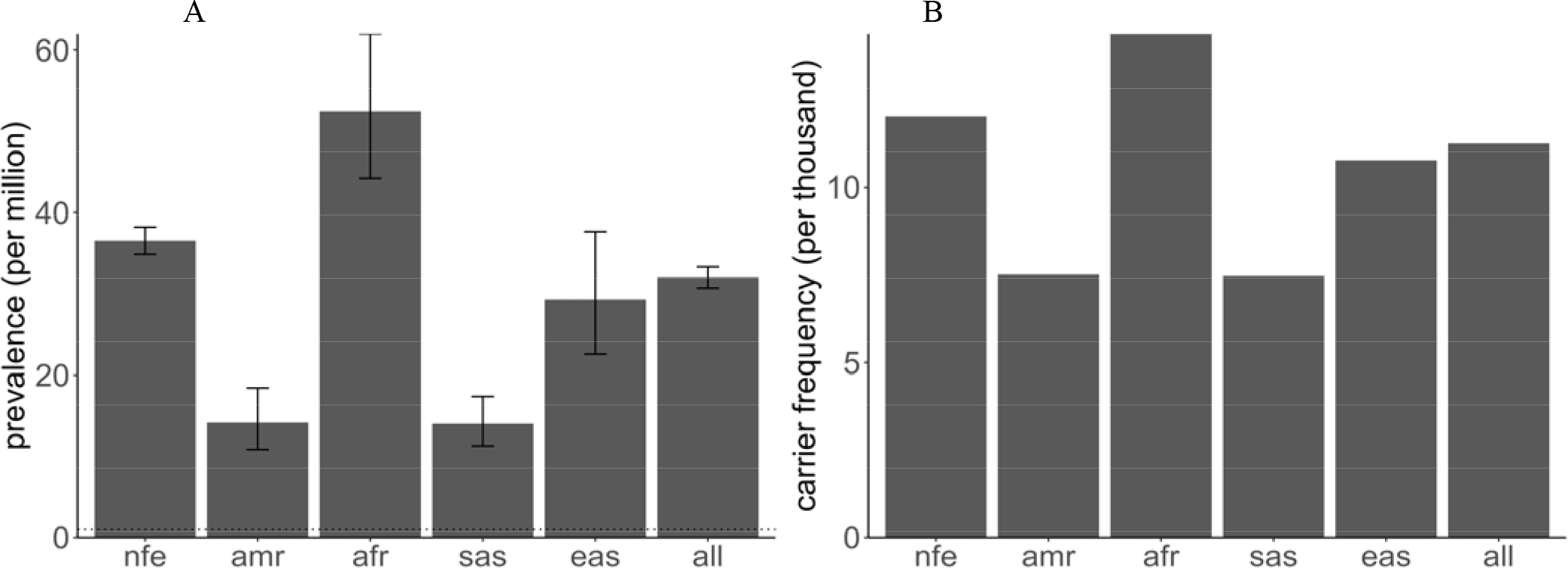
genetic prevalence and carrier frequency of PCD. A. PCD genetic prevalence estimated from gnomAD allele frequencies. B. PCD carrier frequency among diverse population. African/African American (afr), American Admixed/Latino (amr), East Asian (eas), and South Asian (sas).

## Discussion

PCD is a rare autosomal recessive disorder caused by homozygous or compound heterozygous mutations in the solute carrier family 22 member 5 (SLC22A5) gene on chromosome 5q31.1, which encodes organic cation/carnitine transporter 2 (OCTN2) protein. PCD typically manifests in infancy, between the ages of 3 months and 2 years. Infants often present with hypoketotic hypoglycaemia, poor feeding, irritability, lethargy, and hepatomegaly, which is triggered by fasting stress or common illnesses, including gastroenteritis and respiratory tract infections. Approximately half of the patients who present clinically present with muscle hypotonia and progressive childhood cardiomyopathy, which can lead to heart failure. Anemia is occasionally observed in patients with this condition, as carnitine plays a role in red blood cell metabolism. In adults, the presentation is often associated with minor symptoms such as fatigue and decreased stamina. However, dilated cardiomyopathy, arrhythmias, and sudden cardiac death (SCD) have also been reported. Asymptomatic adults have also been described. During pregnancy, minor symptoms as well as cardiac arrhythmias can worsen.

The prevalence is uncertain and varies according to ethnicity. The estimated prevalence is 1/20,000 to 1/70,000 newborns in Europe and the USA, while the estimated incidence in Japan is 1/40,000 births. The prevalence of PCD in the Faroe Islands is the highest reported in the world (1/297). In this study, we sought to estimate the prevalence of PCD using 800 k-scale population genome data and to deepen our understanding of SLC22A5 genetic variation. Using gnomAD data, the prevalence is 3.2 per 1 million (1/31,260) in the global population, 3.6 per million (1/27,388) in the European population, 3 per million (1/34,143) in the East Asian population, and 1.4 per million (1/71,269) in the South Asian population, respectively.

Multi-nucleotide variants (MNVs), defined as two or more nearby variants existing on the same haplotype in an individual, are a clinically and biologically significant consequence of genetic variation [50]. Previously, three MNVs in the SLC22A5 gene have been reported. 3633 of 10583 missense variants were SNVs, but 6950 missense variants were MNVs.

In addition, recent efforts in newborn screening (NBS) have highlighted the importance of second-tier genetic screening for better detection of PCD [3, 14, 51]. Traditional NBS methods, which measure free carnitine (C0) levels, have limitations due to the influence of maternal carnitine levels on the newborn. By incorporating genetic testing, the detection rate of PCD has improved. This combined approach has shown that the actual incidence of PCD may be higher than previously reported.

The prevalence of PCD in the Faroe Islands is the highest reported in the world (1:297) based on 26,462 individuals from the nationwide screening program for primary carnitine deficiency [15]. One study showed a strong association between sudden death and untreated PCD in the Faroe Islands, especially in females [52]. Another study showed PCD in adults can cause serious symptoms, but adult Faroese patients identified through a screening program were predominantly asymptomatic with a normal cardiac structure and function [53]. A 10-year follow-up in the Faroe Islands showed patients with primary carnitine deficiency treated with L-carnitine are alive and doing well more than 10 years after diagnosis [54].

IRIDA is on the first national list of rare diseases issued by China, and the prevalence of IRIDA in China has been estimated [12, 13]. The National Rare Diseases Registry System (NRDRS) has registered IRIDA cases [55]. We demonstrated the power and limitations of the 800-k population genome database to calculate the prevalence of rare diseases. our estimations provide a useful comparison with newborn screening, and for other rare diseases for which screening data are not available, estimations based on genomic data can serve as a valuable reference [16, 17].

In conclusion, through a comprehensive analysis of genetic variation in SLC22A5, we expanded our recognition of disease-causing mutations to 195 variants. These data can be used as a training set for pathogenicity prediction of novel variants and genetic diagnosis of PCD.

## Data Availability

the Science Data Bank (ScienceDB) repository.

https://doi.org/10.57760/sciencedb.09147

## References

1. Magoulas PL, El-Hattab AW: Systemic primary carnitine deficiency: an overview of clinical manifestations, diagnosis, and management. Orphanet J Rare Dis 2012, 7:68.

2. Crefcoeur LL, Visser G, Ferdinandusse S, Wijburg FA, Langeveld M, Sjouke B: Clinical characteristics of primary carnitine deficiency: A structured review using a case-by-case approach. J Inherit Metab Dis 2022, 45(3):386–405.

3. Crefcoeur L, Ferdinandusse S, van der Crabben SN, Dekkers E, Fuchs SA, Huidekoper H, Janssen M, Langendonk J, Maase R, de Sain M et al: Newborn screening for primary carnitine deficiency: who will benefit? - a retrospective cohort study. J Med Genet 2023.

4. Madsen KL, Preisler N, Rasmussen J, Hedermann G, Olesen JH, Lund AM, Vissing J: L-Carnitine Improves Skeletal Muscle Fat Oxidation in Primary Carnitine Deficiency. J Clin Endocrinol Metab 2018, 103(12):4580–4588.

5. Wu X, Prasad PD, Leibach FH, Ganapathy V: cDNA sequence, transport function, and genomic organization of human OCTN2, a new member of the organic cation transporter family. Biochem Biophys Res Commun 1998, 246(3):589–595.

6. Shoji Y, Koizumi A, Kayo T, Ohata T, Takahashi T, Harada K, Takada G: Evidence for linkage of human primary systemic carnitine deficiency with D5S436: a novel gene locus on chromosome 5q. Am J Hum Genet 1998, 63(1):101–108.

7. Nezu J, Tamai I, Oku A, Ohashi R, Yabuuchi H, Hashimoto N, Nikaido H, Sai Y, Koizumi A, Shoji Y et al: Primary systemic carnitine deficiency is caused by mutations in a gene encoding sodium ion-dependent carnitine transporter. Nat Genet 1999, 21(1):91–94.

8. Tamai I, Ohashi R, Nezu J, Yabuuchi H, Oku A, Shimane M, Sai Y, Tsuji A: Molecular and functional identification of sodium ion-dependent, high affinity human carnitine transporter OCTN2. J Biol Chem 1998, 273(32):20378–20382.

9. Wang Y, Ye J, Ganapathy V, Longo N: Mutations in the organic cation/carnitine transporter OCTN2 in primary carnitine deficiency. Proc Natl Acad Sci U S A 1999, 96(5):2356–2360.

10. Tang NL, Ganapathy V, Wu X, Hui J, Seth P, Yuen PM, Wanders RJ, Fok TF, Hjelm NM: Mutations of OCTN2, an organic cation/carnitine transporter, lead to deficient cellular carnitine uptake in primary carnitine deficiency. Hum Mol Genet 1999, 8(4):655–660.

11. Koizumi A, Nozaki J, Ohura T, Kayo T, Wada Y, Nezu J, Ohashi R, Tamai I, Shoji Y, Takada G et al: Genetic epidemiology of the carnitine transporter OCTN2 gene in a Japanese population and phenotypic characterization in Japanese pedigrees with primary systemic carnitine deficiency. Hum Mol Genet 1999, 8(12):2247–2254.

12. Lin Y, Xu H, Zhou D, Hu Z, Zhang C, Hu L, Zhang Y, Zhu L, Lu B, Zhang T et al: Screening 3.4 million newborns for primary carnitine deficiency in Zhejiang Province, China. Clin Chim Acta 2020, 507:199–204.

13. Lin W, Wang K, Zheng Z, Chen Y, Fu C, Lin Y, Chen D: Newborn screening for primary carnitine deficiency in Quanzhou, China. Clin Chim Acta 2021, 512:166–171.

14. Lin Y, Lin B, Chen Y, Zheng Z, Fu Q, Lin W, Zhang W: Biochemical and genetic characteristics of patients with primary carnitine deficiency identified through newborn screening. Orphanet J Rare Dis 2021, 16(1):503.

15. Rasmussen J, Nielsen OW, Janzen N, Duno M, Gislason H, Køber L, Steuerwald U, Lund AM: Carnitine levels in 26,462 individuals from the nationwide screening program for primary carnitine deficiency in the Faroe Islands. J Inherit Metab Dis 2014, 37(2):215–222.

16. Zhao T, Fan S, Sun L: The global carrier frequency and genetic prevalence of Upshaw-Schulman syndrome. BMC Genom Data 2021, 22(1):50.

17. Fan S, Zhao T, Sun L: The global prevalence and ethnic heterogeneity of iron-refractory iron deficiency anaemia. Orphanet J Rare Dis 2023, 18(1):2.

18. Morales J, Pujar S, Loveland JE, Astashyn A, Bennett R, Berry A, Cox E, Davidson C, Ermolaeva O, Farrell CM et al: A joint NCBI and EMBL-EBI transcript set for clinical genomics and research. Nature 2022, 604(7905):310-315.

19. den Dunnen JT, Dalgleish R, Maglott DR, Hart RK, Greenblatt MS, McGowan-Jordan J, Roux A-F, Smith T, Antonarakis SE, Taschner PEM: HGVS Recommendations for the Description of Sequence Variants: 2016 Update. Human Mutation 2016, 37(6):564–569.

20. Frazer J, Notin P, Dias M, Gomez A, Min JK, Brock K, Gal Y, Marks DS: Disease variant prediction with deep generative models of evolutionary data. Nature 2021, 599(7883):91-95.

21. Cheng J, Novati G, Pan J, Bycroft C, Žemgulytė A, Applebaum T, Pritzel A, Wong LH, Zielinski M, Sargeant T, et al: Accurate proteome-wide missense variant effect prediction with AlphaMissense. Science 2023, 381(6664):eadg7492.

22. Brandes N, Goldman G, Wang CH, Ye CJ, Ntranos V: Genome-wide prediction of disease variant effects with a deep protein language model. Nature Genetics 2023, 55(9):1512–1522.

23. Schubach M, Maass T, Nazaretyan L, Röner S, Kircher M: CADD v1.7: using protein language models, regulatory CNNs and other nucleotide-level scores to improve genome-wide variant predictions. Nucleic Acids Research 2024, 52(D1):D1143–D1154.

24. Gao H, Hamp T, Ede J, Schraiber JG, McRae J, Singer-Berk M, Yang Y, Dietrich ASD, Fiziev PP, Kuderna LFK et al: The landscape of tolerated genetic variation in humans and primates. Science 2023, 380(6648):eabn8153.

25. Allot A, Wei CH, Phan L, Hefferon T, Landrum M, Rehm HL, Lu Z: Tracking genetic variants in the biomedical literature using LitVar 2.0. Nat Genet 2023, 55(6):901–903.

26. Harrison PW, Amode MR, Austine-Orimoloye O, Azov Andrey G, Barba M, Barnes I, Becker A, Bennett R, Berry A, Bhai J, et al: Ensembl 2024. Nucleic Acids Research 2023, 52(D1):D891-D899.

27. Sayers Eric W, Beck J, Bolton Evan E, Brister J R, Chan J, Comeau Donald C, Connor R, DiCuccio M, Farrell Catherine M, Feldgarden M et al: Database resources of the National Center for Biotechnology Information. Nucleic Acids Research 2023, 52(D1):D33–D43.

28. Jaganathan K, Kyriazopoulou Panagiotopoulou S, McRae JF, Darbandi SF, Knowles D, Li YI, Kosmicki JA, Arbelaez J, Cui W, Schwartz GB et al: Predicting Splicing from Primary Sequence with Deep Learning. Cell 2019, 176(3):535–548.e524.

29. Zeng T, Li YI: Predicting RNA splicing from DNA sequence using Pangolin. Genome Biology 2022, 23(1):103.

30. Zhang J, Yao Y, He H, Shen J: Clinical Interpretation of Sequence Variants. Curr Protoc Hum Genet 2020, 106(1):e98.

31. Richards S, Aziz N, Bale S, Bick D, Das S, Gastier-Foster J, Grody WW, Hegde M, Lyon E, Spector E et al: Standards and guidelines for the interpretation of sequence variants: a joint consensus recommendation of the American College of Medical Genetics and Genomics and the Association for Molecular Pathology. Genet Med 2015, 17(5):405–424.

32. Tavtigian SV, Greenblatt MS, Harrison SM, Nussbaum RL, Prabhu SA, Boucher KM, Biesecker LG: Modeling the ACMG/AMP variant classification guidelines as a Bayesian classification framework. Genet Med 2018, 20(9):1054–1060.

33. Biesecker LG, Harrison SM, ClinGen Sequence Variant Interpretation Working G: The ACMG/AMP reputable source criteria for the interpretation of sequence variants. Genet Med 2018, 20(12):1687–1688.

34. Ghosh R, Harrison SM, Rehm HL, Plon SE, Biesecker LG, ClinGen Sequence Variant Interpretation Working G: Updated recommendation for the benign stand-alone ACMG/AMP criterion. Hum Mutat 2018, 39(11):1525–1530.

35. Abou Tayoun AN, Pesaran T, DiStefano MT, Oza A, Rehm HL, Biesecker LG, Harrison SM, ClinGen Sequence Variant Interpretation Working G: Recommendations for interpreting the loss of function PVS1 ACMG/AMP variant criterion. Hum Mutat 2018, 39(11):1517–1524.

36. Brnich SE, Abou Tayoun AN, Couch FJ, Cutting GR, Greenblatt MS, Heinen CD, Kanavy DM, Luo X, McNulty SM, Starita LM et al: Recommendations for application of the functional evidence PS3/BS3 criterion using the ACMG/AMP sequence variant interpretation framework. Genome Med 2019, 12(1):3.

37. Pejaver V, Byrne AB, Feng BJ, Pagel KA, Mooney SD, Karchin R, O’Donnell-Luria A, Harrison SM, Tavtigian SV, Greenblatt MS et al: Calibration of computational tools for missense variant pathogenicity classification and ClinGen recommendations for PP3/BP4 criteria. Am J Hum Genet 2022, 109(12):2163–2177.

38. Walker LC, Hoya M, Wiggins GAR, Lindy A, Vincent LM, Parsons MT, Canson DM, Bis-Brewer D, Cass A, Tchourbanov A et al: Using the ACMG/AMP framework to capture evidence related to predicted and observed impact on splicing: Recommendations from the ClinGen SVI Splicing Subgroup. Am J Hum Genet 2023, 110(7):1046–1067.

39. Preston CG, Wright MW, Madhavrao R, Harrison SM, Goldstein JL, Luo X, Wand H, Wulf B, Cheung G, Mandell ME et al: ClinGen Variant Curation Interface: a variant classification platform for the application of evidence criteria from ACMG/AMP guidelines. Genome Med 2022, 14(1):6.

40. Koleske ML, McInnes G, Brown JEH, Thomas N, Hutchinson K, Chin MY, Koehl A, Arkin MR, Schlessinger A, Gallagher RC et al: Functional genomics of OCTN2 variants informs protein-specific variant effect predictor for Carnitine Transporter Deficiency. Proc Natl Acad Sci U S A 2022, 119(46):e2210247119.

41. Frigeni M, Balakrishnan B, Yin X, Calderon FRO, Mao R, Pasquali M, Longo N: Functional and molecular studies in primary carnitine deficiency. Hum Mutat 2017, 38(12):1684–1699.

42. Li FY, El-Hattab AW, Bawle EV, Boles RG, Schmitt ES, Scaglia F, Wong LJ: Molecular spectrum of SLC22A5 (OCTN2) gene mutations detected in 143 subjects evaluated for systemic carnitine deficiency. Hum Mutat 2010, 31(8):E1632–1651.

43. Wang Y, Korman SH, Ye J, Gargus JJ, Gutman A, Taroni F, Garavaglia B, Longo N: Phenotype and genotype variation in primary carnitine deficiency. Genet Med 2001, 3(6):387–392.

44. Amat di San Filippo C, Pasquali M, Longo N: Pharmacological rescue of carnitine transport in primary carnitine deficiency. Hum Mutat 2006, 27(6):513-523.

45. El-Hattab AW, Li FY, Shen J, Powell BR, Bawle EV, Adams DJ, Wahl E, Kobori JA, Graham B, Scaglia F et al: Maternal systemic primary carnitine deficiency uncovered by newborn screening: clinical, biochemical, and molecular aspects. Genet Med 2010, 12(1):19–24.

46. Rose EC, di San Filippo CA, Ndukwe Erlingsson UC, Ardon O, Pasquali M, Longo N: Genotype-phenotype correlation in primary carnitine deficiency. Hum Mutat 2012, 33(1):118-123.

47. Verbeeten KC, Lamhonwah AM, Bulman D, Faghfoury H, Chakraborty P, Tein I, Geraghty MT: Carnitine uptake defect due to a 5’UTR mutation in a pedigree with false positives and false negatives on Newborn screening. Mol Genet Metab 2020, 129(3):213–218.

48. Ferdinandusse S, Te Brinke H, Ruiter JPN, Haasjes J, Oostheim W, van Lenthe H, L IJ, Ebberink MS, Wanders RJA, Vaz FM et al: A mutation creating an upstream translation initiation codon in SLC22A5 5’UTR is a frequent cause of primary carnitine deficiency. Hum Mutat 2019, 40(10):1899–1904.

49. Lamhonwah AM, Olpin SE, Pollitt RJ, Vianey-Saban C, Divry P, Guffon N, Besley GT, Onizuka R, De Meirleir LJ, Cvitanovic-Sojat L et al: Novel OCTN2 mutations: no genotype-phenotype correlations: early carnitine therapy prevents cardiomyopathy. Am J Med Genet 2002, 111(3):271–284.

50. Wang Q, Pierce-Hoffman E, Cummings BB, Alföldi J, Francioli LC, Gauthier LD, Hill AJ, O’Donnell-Luria AH, Armean IM, Banks E et al: Landscape of multi-nucleotide variants in 125,748 human exomes and 15,708 genomes. Nature Communications 2020, 11(1):2539.

51. Lin Y, Zhang W, Huang C, Lin C, Lin W, Peng W, Fu Q, Chen D: Increased detection of primary carnitine deficiency through second-tier newborn genetic screening. Orphanet J Rare Dis 2021, 16(1):149.

52. Rasmussen J, Dunø M, Lund AM, Steuerwald U, Hansen SH, Joensen HD, Køber L, Nielsen OW: Increased risk of sudden death in untreated primary carnitine deficiency. J Inherit Metab Dis 2020, 43(2):290–296.

53. Rasmussen J, Køber L, Lund AM, Nielsen OW: Primary Carnitine deficiency in the Faroe Islands: health and cardiac status in 76 adult patients diagnosed by screening. J Inherit Metab Dis 2014, 37(2):223–230.

54. Abrahamsen RK, Lund AM, Rasmussen J: Patients with primary carnitine deficiency treated with L-carnitine are alive and doing well—A 10-year follow-up in the Faroe Islands. JIMD Reports 2023, 64(6):453–459.

55. Guo J, Liu P, Chen L, Lv H, Li J, Yu W, Xu K, Zhu Y, Wu Z, Tian Z et al: National Rare Diseases Registry System (NRDRS): China’s first nation-wide rare diseases demographic analyses. Orphanet J Rare Dis 2021, 16(1):515.

